# Comparison of collection methods for *Phlebotomus argentipes* sand flies to use in a molecular xenomonitoring system for the surveillance of visceral leishmaniasis

**DOI:** 10.1101/2023.02.28.23286557

**Authors:** Shannon McIntyre-Nolan, Vijay Kumar, Miguella Mark Carew, Kundan Kumar, Emily Nightingale, Giorgia Dalla Libera Marchiori, Matthew Rogers, Mojca Kristan, Susana Campino, Graham F. Medley, Pradeep Das, Mary Cameron

**Affiliations:** Department of Disease Control, London School of Hygiene & Tropical Medicine, London, UK; Rajendra Memorial Research Institute of Medical Sciences, Patna, India; Department of Global Health and Development, London School of Hygiene & Tropical Medicine, London, UK; Department of Infection Biology, London School of Hygiene & Tropical Medicine, London, UK; Department of Molecular Parasitology, National Institute of Cholera and Enteric Diseases, Kolkata 10, India

**Author notes:** These authors contributed equally to this work (Shannon McIntyre-Nolan, Vijay Kumar). These authors also contributed equally to this work (Pradeep Das, Mary Cameron).

**Keywords:** Molecular xenomonitoring, visceral leishmaniasis, neglected tropical diseases (NTDs), disease surveillance, disease elimination, *Phlebotomus argentipes*, *Leishmania donovani*

## Abstract

**Background:** The kala-azar elimination programme has resulted in a significant reduction in visceral leishmaniasis (VL) cases across the Indian Subcontinent. To detect any resurgence of transmission, a sensitive cost-effective surveillance system is required. Molecular xenomonitoring (MX), detection of pathogen DNA/RNA in vectors, provides a proxy of human infection in the lymphatic filariasis elimination programme. To determine whether MX can be used for VL surveillance in a low transmission setting, large numbers of the sand fly vector *Phlebotomus argentipes* are required. This study will determine the best method for capturing *P. argentipes* females for MX.

**Methodology/Principal Findings:** The field study was performed in two programmatic and two non-programmatic villages in Bihar, India. A total of 48 households (12/village) were recruited. Centers for Disease Control and Prevention light traps (CDC-LTs) were compared with Improved Prokopack (PKP) and mechanical vacuum aspirators (MVA) using standardised methods. Four 12×12 Latin squares, 576 collections, were attempted (12/house, 144/village,192/method). Molecular analyses of collections were conducted to confirm identification of *P. argentipes* and to detect human and *Leishmania* DNA. Operational factors, such as time burden, acceptance to householders and RNA preservation, were also considered. A total of 562 collections (97.7%) were completed with 6,809 sand flies captured. Females comprised 49.0% of captures, of which 1,934 (57.9%) were identified as *P. argentipes*. CDC-LTs collected 4.04 times more *P. argentipes* females than MVA and 3.62 times more than PKP (p<0.0001 for each). Of 21,735 mosquitoes in the same collections, no significant differences between collection methods were observed. CDC-LTs took less time to install and collect than to perform aspirations and their greater yield compensated for increased sorting time. CDC-LTs were favoured by householders.

**Conclusions/Significance:** CDC-LTs are the most useful collection tool of those tested for MX surveillance since they collected higher numbers of *P. argentipes* females without compromising mosquito captures or the preservation of RNA. However, capture rates are still low.

**Author Summary:** Molecular xenomonitoring, screening insects for pathogen DNA/RNA, may be used for surveillance of diseases transmitted by insects. Since the proportion of insects infected with pathogens is very low in areas targeted for disease elimination, large numbers of females need to be screened. We compared three different methods for collecting *Phlebotomus argentipes* sand fly females, the vector of parasites causing the disease visceral leishmaniasis in the Indian subcontinent, to determine which collected the largest number of females. Other factors that may also influence selection of a particular method of collection by a disease control programme, such as the time it takes to collect and sort samples, the acceptance of householders for a particular collection method and whether RNA degradation in insect samples differed between collection methods, were also considered. Centers for Disease Control and Prevention light traps (CDC-LTs) proved to be more useful than two types of aspiration methods for collecting higher numbers of sand fly females and RNA preservation was retained. Furthermore, they took less time to install than to perform aspirations and were favoured by householders. Therefore, CDC-LTs were considered to be the most suitable collection method for molecular xenomonitoring of sand flies in India.

## Introduction

### Visceral Leishmaniasis (VL) Epidemiology and Transmission in India

Across the Indian Subcontinent (ISC) VL, also known as kala-azar, is an anthroponotic disease caused by the parasite *Leishmania donovani*. It is vectored by only one incriminated sand fly species, *Phlebotomus argentipes* (1). In 2005, Bangladesh, India and Nepal accounted for 70% of the global VL burden but, given its epidemiology in the region, together with newer tools for diagnosis and treatment, the prospect of elimination of VL as a public health problem became a feasible goal. Consequently, a memorandum of understanding was signed by the three regional members at the WHO Headquarters in Geneva, Switzerland to eliminate VL as a public health problem with an agreed target to reduce incidence to below 1 case/10,000 population at the country’s appropriate administration level, upazila, block, or sub-district, by 2015 (2).

Since the implementation of the elimination strategy, consisting of rapid case detection, treatment of VL cases and vector control using indoor residual spraying (IRS), and the accelerated plan for elimination practiced by the National Vector Borne Disease Control Programme (NVBDCP) in India (3), the number of reported VL cases in Bangladesh, India, and Nepal has declined sharply, from 41,158 in 2005 to 4,692 in 2018, and the global burden of VL in the ISC has reduced from 70% in 2005 to an unprecedented low of 27% (4). Bangladesh and Nepal have reached the elimination target in all endemic areas, while India has reached the target in 714/751 (95%) endemic blocks (i.e. those deemed to have on-going transmission). In 2019, all 37 blocks that were above target were located in two of four VL endemic states: Bihar and Jharkhand (21 and 16 blocks, respectively) (5).

This progress is welcomed, but it brings enormous challenges to the National Kala-azar Elimination Programme since, as VL cases become scarce, it is crucial to have sensitive and cost-effective surveillance systems in place to determine whether elimination targets have been met and sustain control to prevent resurgence. Molecular xenomonitoring (MX) is a vector-based pathogen surveillance system, detecting pathogen DNA/RNA in a vector as a proxy of human infection, that may serve as a useful alternative to monitoring human infection and residual transmission in this low-transmission context (6).

### Molecular Xenomonitoring (MX)

Much of the previous literature on MX concerns its use in lymphatic filariasis (LF) elimination programmes, where endpoints of transmission have been established through several decades of research, to determine whether transmission is still occurring after mass drug administration (MDA) (6). Establishing the appropriate sample size for a given MX system depends on its goal. For elimination, endpoints of transmission are used to determine if elimination is nearing, has been achieved, or has been sustained (6). Sample size calculations depend on estimates from existing literature on vector abundance and infection prevalence (ideally from the same, nearby, or characteristically similar sites). In areas where elimination activities are underway, and human and vector infection prevalence have or can be assumed to have decreased, significantly more vectors will be required to detect low infection levels than if infection levels were high (6). For example, for LF elimination surveillance, the numbers of mosquitoes required to obtain accurate filarial prevalence rates in different geographical settings post-MDA depends on vector efficiency and may be as high as 5,000 - 7,500 *Culex quinquefasciatus*, 2,500 *Anopheles* and 22,000 *Aedes* females (7).

Consequently, several studies were performed to obtain the large numbers of mosquitoes required and to gain an understanding of their behaviour since this influences the ideal tool and method of collection (6). In general, for *Cx. quinquefasciatus*, Centers for Disease Control (CDC) and Prevention gravid traps (GTs) baited with odorous infusions, exploiting oviposition behaviour, are placed outside of houses and are the preferred method for collecting gravid adult females. However, a comparison of collection methods in Brazil found that large battery-powered aspirators, exploiting host-seeking behaviour, collected 38 times more blood-fed and 5 times more gravid *Cx. quinquefasciatus* than CDC light traps (LTs) placed inside houses (8). Furthermore, residents preferred aspirators over fixed battery or AC powered traps (e.g., CDC-LTs) due to lower risk of battery theft, power cuts, and the nuisance of light and noise in the bedroom at night (8). Similarly, for arbovirus surveillance, common methods for collecting mosquitoes have known advantages and disadvantages (9). However, the optimal collection method to use for *P. argentipes*, to develop an MX system for *L. donovani* DNA surveillance, has not been explored.

### Rationale for MX system for VL surveillance in India

A better understanding of the transmission dynamics of VL, in particular of how rates of infection in humans and sand flies vary as functions of each other, is required to guide VL elimination efforts and ensure sustained elimination in the ISC. By collecting contemporary entomological and human data in the same geographical locations, more precise epidemiological models can be produced allowing MX endpoints of VL transmission to be determined similar to those obtained for LF. For example, the entomological inoculation rate (EIR), a product of the human bloodmeal index (HBI) and the proportion of vectors that are infective, a useful proxy for human infection in malaria control programmes, is unknown for VL (10). In order to calculate the EIR for VL in the ISC, a suitable collection method to obtain large numbers of *P. argentipes* females is required.

Systematic longitudinal sampling of *P. argentipes* populations to monitor the impact of IRS on their abundance, HBI, and the presence of *L. donovani* infection is not performed routinely in India, Bangladesh or Nepal. Nevertheless, several research studies have captured *P. argentipes* for different purposes, and the most common methods use either mouth aspirations or CDC-LTs placed in human dwellings or in cattle sheds (11). Specific details concerning trap placement and measures taken to standardize collections, which is a particularly important concern when mouth aspirations are used, are often missing from surveys making comparisons between them difficult. Furthermore, collections of blood-fed females are often limited, which adds to the potential bias arising through sampling procedures and can lead to data misinterpretation (12). More recently, studies in Bihar have improved the standardization of collections using CDC-LTs and found that *P. argentipes* feed preferentially on humans and, a higher proportion of human-fed *P. argentipes* are found in cattle enclosures (55%) compared with houses (31%) (13, 14).

The first study to examine natural infection rates of *P. argentipes* with *L. donovani* in Bihar collected a total of 14,585 sand flies using CDC miniature light traps and mouth aspirators in the Muzaffarpur district (15). Of these, a subset of 449 *P. argentipes* females were divided into pools for molecular detection of the 18S rRNA gene using PCR, but the overall prevalence of infection in *P. argentipes* for *L. donovani* DNA was over estimated as 4.90-17.37% because the number of individual sand flies that may be positive in a single pool was not taken into account (15). Subsequently, more reliable estimates were obtained using individual sand flies and infection rates of *P. argentipes* with *L. donovani* DNA were found to vary according to season: 1.0% (4/384) in summer, 0.9% (5/591) in the rainy season and 2.8% (12/422) in winter (16). However, this study was performed over a single year, and morphological identifications were not confirmed using molecular methods, so further work is required to examine seasonal trends. Since then, studies have collected *P. argentipes* using CDC-LTs and/or mouth aspirators (14, 17) but no natural infection rates of *P. argentipes* for *L. donovani* DNA were published in the last seven years prior to the commencement of the present study.

The primary aim of the present study is to compare collection methods to determine which method collects the largest number of *P. argentipes* females for use in subsequent epidemiological studies where the EIR and endpoints for transmission will be calculated. CDC gravid traps (CDC-GTs), baited with water rather than organic infusions for indoor use, were a method considered for inclusion because of their widescale deployment in LF surveillance, but they were not selected in the present trial since it was previously shown that collections of sand flies (including live as well as dead females of different physiological stages), were higher in CDC-LTs than in CDC-GTs in Odisha, India (18). Therefore, CDC-LTs, often considered a ‘gold standard’ for *P. argentipes* (11), will be compared with two different types of battery-operated aspirators that were shown to collect more mosquitoes resting indoors than CDC-LTs or CDC backpack aspirators in earlier trials (8, 19). Although *P. argentipes* is the primary species of interest, incidental captures of mosquito females will also be analysed. In addition to entomological indicators, including human bloodmeal analysis and *L. donovani* infection rates, operational factors relating to the time burden of different collection methods, and their acceptance to householders, will be examined.

Another important consideration when selecting an appropriate collection technique for MX is whether pathogen RNA is preserved in the vector during collection and subsequent transportation prior to storage at -80°C. RNA detection is required for screening vectors for the infective stages of parasites, the metacyclic stages in the case of *Leishmania*, to measure EIR. To preserve the integrity of samples, a cold chain is required during transport of samples to the laboratory where they can be placed in RNA preservative media and frozen (9, 20). However, it is possible that the different conditions experienced by sand flies between collection methods, where sand flies are placed in a cool bag immediately after aspiration versus being trapped in a CDC-LT for possibly 12 or more hours before being placed in a cool bag, may compromise RNA preservation which will be addressed through a laboratory experiment simulating field conditions.

## Methods

### Study Sites

The field study was performed in Bihar, the state in India with the highest incidence of VL. Two villages each in Nalanda (Ruchanpura, also known as Kosiawan, in Ekangarsarai block and Dharampur in Thartari block) and Saran (Bishambharpur and Rampur Jagdish, both in Dariapur block) districts were selected for sand fly collection (Fig 1). Nalanda is located south of the Ganges River (25° 12′ 0″ N, 85° 31′ 12″ E); the Ganges River forms the southern border of Saran (25° 55′ 0″ N, 84° 45′ 0″ E). Travel distance to the villages from Rajendra Memorial Research Institute of Medical Sciences (RMRIMS), where the field team were based, ranged from 41-50 kilometres (per Google Maps) and travel time was estimated to be between 1.5 to 2.5 hours depending on traffic conditions.

**Fig 1.**
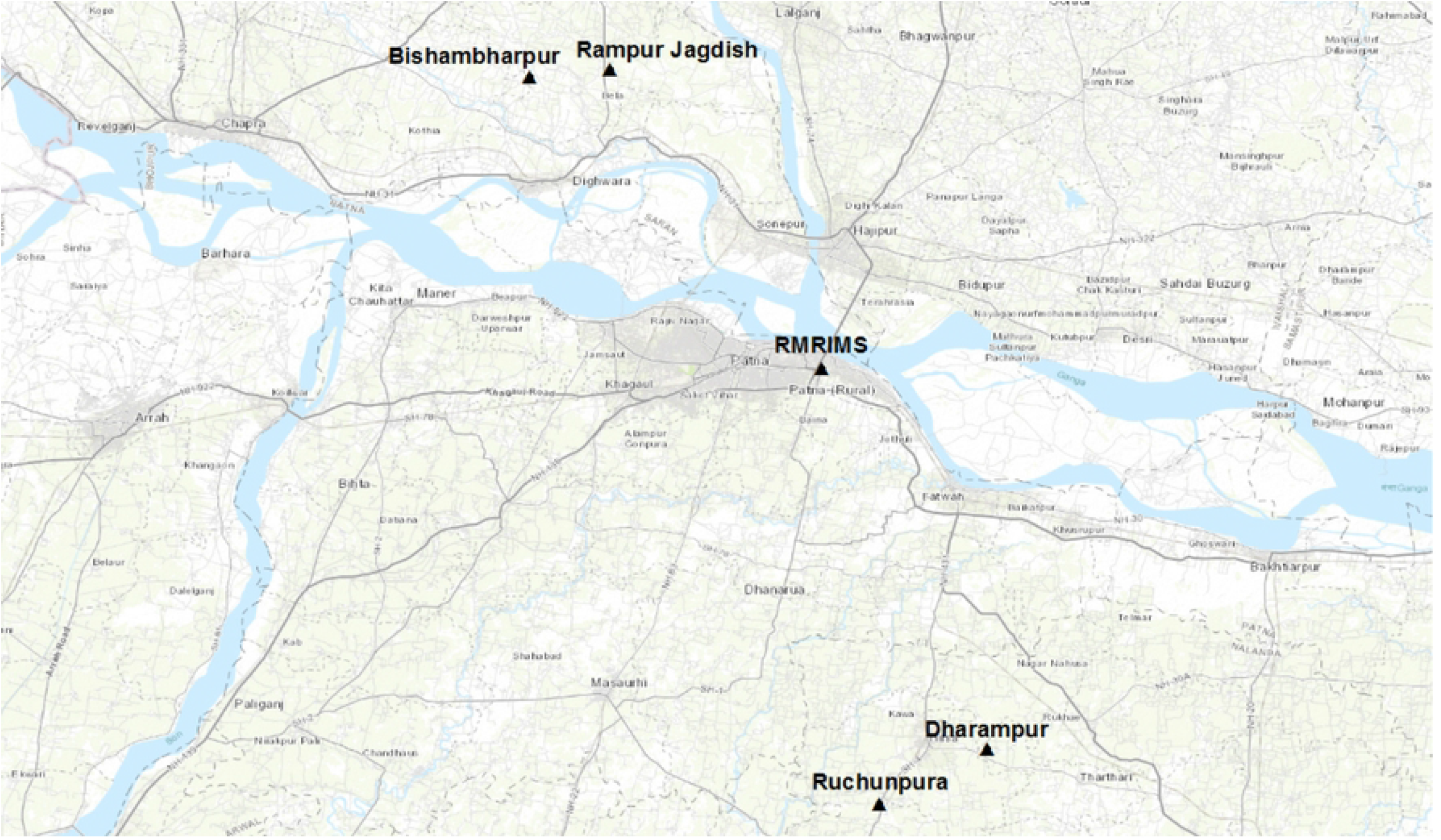
Location of study villages and RMRIMS in Bihar, India.

Nalanda villages were defined as non-programmatic villages, i.e. at the time of sampling they met and sustained the Government of India’s elimination target of <1 VL case reported/10,000 population at the block level over a three-year period, and no IRS or other vector control activities occurred in the five years preceding this study. Saran villages were defined as programmatic villages, meaning that they were above the elimination target at the time of sampling and should receive IRS twice during the calendar year.

Household sampling sites were identified in each village based on pre-study vector surveys. The final selection of households included human-only and combined human-cattle dwellings. Replacement was needed for three households over the course of the study.

### Collection Method Comparison (CMC)

#### Sample Size Calculation

Prior to the start of the study, a sample size calculation using data collected during a previous trial conducted in Bihar, where a mean of 1.9776 *P. argentipes* females, with a standard deviation of 3.2512, were collected with CDC-LTs per trap night (21), determined that 166 trap nights/collection method was sufficient to detect a difference of one sand fly in the mean number of *P. argentipes* females collected by each of the three methods (where *α =* 0.05; ß = 0.20). To determine the best practice for sampling female *P. argentipes* sand flies, CDC-LTs (John Hock) were compared with improved Prokopack aspirators (PKP) (John Hock) and mechanical vacuum aspirators (MVA) (Horst Armadilhas).

#### Design

A total of 48 households (12 from each of the four villages) were recruited for regular indoor sampling four days a week over 12 weeks from 25 June to 14 September 2018. Using a Latin Square design balanced for carryover effects, the three collection methods were rotated through each household with four 12×12 Latin Squares (rounds) over the study period. A total of 576 collection events were expected (288 events per district, 144 events per village, 192 collection events per collection method, and 12 collection events per household).

#### CDC-LT Protocol

When allocated, a CDC-LT was installed in each of participating households and turned on between 18.00 hours and 06.00 hours based on the day of treatment according to the Latin square design. Batteries (6V 12 amp) were charged at RMRIMS prior to installation. CDC-LTs were hung in the room where most household residents slept at a distance 15cm from the wall of the house with the base of the collection pot 5-10cm from the floor. A member of the field team connected one receptor of the CDC-LT to the appropriate terminal of the battery and instructed a household member on how to connect the second receptor between 18.00 and 19.00 on the installation day.

Householders were contacted via mobile phone near this time to ensure that traps were turned on for the expected duration. The field team returned the next morning to collect the CDC-LTs.

#### PKP and MVA Protocols

At the time of recruitment, the internal area of the room where PKP and MVA aspirations were to be performed was measured. The amount of time spent aspirating each room was calculated based on this room-specific measurement and an aspiration speed of approximately of 1m² per 30 seconds. Two PKP (12V 12 amp) and two MVA (12 V 5 amp) batteries were charged overnight prior to use the following morning. Aspirations began between 07:00 and 08:00 and continued until all houses assigned to these collection methods on the day were completed.

Two field workers were needed to perform PKP and MVA aspirations: one to operate the PKP or MVA and another to operate a timer and remove objects obstructing the aspiration path. Aspirations always commenced on the wall immediately to the left of the doorway, on entering the room, and were continued clockwise around the room. Space below furniture, such as beds and tables, was also aspirated. In instances where the operator completed all walls before the allocated time had expired, a second round commenced until the calculated aspiration time was reached. Walls were aspirated to a height of up to approximately seven feet from the floor or to the ceiling, whichever was lower. Aspiration occurred left to right across the wall, at a distance of approximately 1cm from the surface, and gradually shifted downwards covering the entire area to the bottom of the wall. Contents of collection tubes were observed after aspiration for the presence of spiders, which if found were killed due to the risk of predation of sand flies.

#### Post-Collection Storage, Transport, and Processing

Specimens from all three collection methods were transported in holding containers placed in insulated coolers, containing ice packs, to RMRIMS. Due to the limited number of capture bags available for the MVAs, and collection containers for the PKP, aspirations were transferred to universal tubes fitted with fine mesh and secured by a screw cap with a hole in it for transfer of specimens using a small, handheld battery-operated aspirator. Holes were plugged with moist cotton wool to retain humidity and were secured with autoclave tape immediately after transfer for transport and storage. Universal tubes and CDC-LT collection pots were all stored at -20°C to freeze kill specimens prior to processing. Tubes and collection pots were labelled with the date of collection and a unique household number that also conferred geographic (district and village) information.

Collections were sorted via microscopy. Sand flies were separated from other collected arthropods and enumerated by sex and physiological status. Female and male sand flies were placed in separate microcentrifuge tubes containing RNALater™ with a maximum of 25 sand flies per tube. Specimens were stored in cryoboxes at -20°C.

#### Data Collection and Analysis

Data for each collection event (date of collection, district, village, sex and physiological state of sand flies) were recorded in a Microsoft Excel database. Descriptive statistics included analysis of total sand fly and total female mosquito data by collection method, district and village. The number of female *P. argentipes* per collection was modelled using negative binomial regression, with independent and identically distributed random effects to account for clustering induced by multiple collections per household. Data for the fourth round of collections were excluded from statistical analyses since specimens were lost during storage and species data were not obtainable. The proportion of human blood-fed females out of all female *P. argentipes* caught was compared between collection methods using logistic regression, also accounting for household clustering. All statistical analyses were performed using R (version 3.6.3 (2020-02-29)).

### Molecular Analyses

#### DNA Extraction

DNA was extracted from all female sand flies using the Qiagen DNeasy Blood and Tissue Kit per manufacturer’s protocol. DNA samples were stored at -20°C. Molecular analyses were performed to confirm microscopic species identification of *P. argentipes*, for human DNA detection and *L. donovani* DNA detection in *P. argentipes* females.

#### Species Identification via PCR

All female sand flies were identified using a PCR-RFLP protocol targeting the 18S rRNA coding genes for phlebotomine sandflies (22), which was adapted as described below. To differentiate *P. argentipes* from *P. papatasi*, and members of the *Sergentomyia babu* complex, PCR products underwent restriction enzyme digestion using *HinfI* and *HpalI* separately for one hour at 37°C. The total volume per reaction was 25µl, which include 12.5µl of Taq PCR master mix (Qiagen), 1.25 µl for each 10pmol/ µl primer (Forward 18S primer: 5’-TAGTGAAACCGCAAAAGGCTCAG-3’; Reverse 18S primer: 5’-CTCGGATGTGAGTCCTGTATT GT-3’) and 10 µl of DNA sample. Digested products were run on a 2% agarose gel followed by ethidium bromide staining to detect bands of appropriate sizes corresponding to the three sandfly species.

#### Human DNA analysis

Female sand flies identified as *P. argentipes* by PCR-RFLP were analysed for the presence of human DNA using a qPCR protocol with primers targeting the cytochrome b (cytb) gene as described previously (23). Quantitative detection of human DNA was performed on an Applied Biosystems 7500 Fast Real-Time PCR System according to the KAPA SYBR® FAST Universal Master Mix recommended protocol. For standard curves, human DNA obtained from purchased donor blood (Cambridge Bioscience) was serially diluted to provide a range of 1-0.0001ng/µl. DNA free water was used as No Template Control (NTC) in each assay. A total of 5µl of DNA was used from each female *P. argentipes*. Samples were considered positive if Ct values were lower than the lower limit of detection of the assay (Ct< 30). All samples and controls were run in duplicate.

#### Leishmania donovani DNA analysis

Female sand flies identified as *P. argentipes* by PCR-RFLP were analysed for the presence of *L. donovani* DNA using a qPCR protocol with Taqman primers and probes as previously described and validated (24, 25). Quantitative detection of *Leishmania* DNA was performed on an Applied Biosystems 7500 Fast Real-Time PCR System using the conditions recommended by the KAPA Probe® FAST Universal kit protocol. For standard curves, *L. donovani* DNA from the reference strain DD8 was serially diluted to provide a range of 1-10^−5^ ng/µl. DNA from non-infected and experimentally infected sand flies from the LSHTM colony was used as negative and positive control, respectively. DNA free water was used as NTC in each assay. A total of 10µl of DNA was used from each female *P. argentipes*. Samples were considered positive if Ct values were lower than the limit of detection of the assay (Ct< 31). All samples and controls were run in duplicate.

#### Simulation of Leishmania donovani RNA degradation

In order to simulate the conditions that infective sand flies may experience during collections, sand flies were experimentally infected with *L. donovani* at high infection rates of 3×10^6^ amastigotes/ml heat-inactivated human blood through a chick skin (equivalent to 3000 amastigotes/sandfly) and maintained at 28 °C, 80% relative humidity on 10% (w/v) sucrose for 8 days to allow the parasites to colonise the anterior midgut and produce metacyclic promastigotes, the infectious forms. Sand flies were then killed and kept at different treatment groups simulating the conditions they would experience either during aspiration or collection by a CDC-LT and prior to transportation to the laboratory for RNA extraction and analysis using *Leishmania*-specific ssrRNA primers.

The four treatment groups considered relevant for aspirations, where sandflies are placed in a cool bag shortly after collection at ambient temperatures of up to 40°C and then transported to the laboratory, were RNA extraction at: T0 (immediately after killing), T1 (after 30 mins, kept at 40°C), T2 (after 3 h, kept in a cool bag) and T3 (after 7 h, kept in a cool bag). For CDC-LTs, where sandflies are collected overnight prior to transportation to the laboratory, T1 was revised to after 13-16 h, kept at 40°C, as a ‘worst case scenario’ for possible RNA degradation, and this treatment was repeated using sand flies infected with a low rate considered to be more representative to field conditions (equivalent to <100 amastigotes/sandfly).

### Operational considerations

#### Time burden of sampling

To assess logistical constraints when using CDC-LT, MVA or PKP in a field setting, 24 additional trap-nights were attempted (8 per method) in a supplement study performed in the Saran villages in February 2019. The total time burden required to carry out collections for each method was recorded to the nearest minute for each household. For CDC-LTs, this was measured as the installation time burden. Additionally, the total time required to sort the samples in each collection pot according to the protocols implemented during the trapping comparison study was recorded to the nearest second.

#### Householder acceptance

To understand acceptance of and preferences for the three collections methods, all 48 households were surveyed to determine their preferred collection method, the rationale for their choice, and other considerations for the three methods. The investigators asked householders the following questions:

1. Do you prefer collections performed by: CDC light traps, mechanical vacuum aspirators, Prokopack aspirators or have no preference?
2. What is your reason for the answer given to question 1?
3. Do you have any additional comments/complaints relating to any of the collection methods?

## Results

### CMC descriptive analysis of global data set

#### *Collection* site

Of a total of 576 attempted individual collection events, 562 (97.7%) were completed successfully. Battery failure (for CDC-LTs left overnight) and inability to enter a household were the main reasons for unsuccessful collection events. Failures occurred across the four study villages, with one household having two failed collection events. A total of 6,809 sand flies were collected over the study period. At least one sand fly was collected from each household with an outlier of 396 total sand flies reported from one household in Ruchunpura (Nalanda) over the study period. Over 82% of collection events yielded at least one sand fly (465/562).

Fig 2 shows the number of specimens by collection site. Almost half of the sand flies collected were female (49.0%, n=3,339). Of 3,232 female sand flies identified to species by PCR, 1,934 (59.8%) were *P. argentipes. Sergentomyia babu* accounted for 37.0% (n=1,195) of females, and *P. papatasi* accounted for 3.2% (n=103). The remaining 107 samples were unable to be identified to species. Although the proportion of total female sand flies collected in the two districts was roughly equal, the species composition differed. More *P. argentipes* females (n = 1,024, 52.9% versus n = 910, 47.1%) and *P. papatasi* females (n = 102, 99.0% versus n = 1, 1.0%) were collected in Saran than in Nalanda, respectively. Most of the *P. papatasi* females were collected from Rampur Jagdish (n = 95, 92.2%). Of 21,735 mosquitoes collected, 8,174 (37.6%) were female. Conversely, more mosquito females were collected in Nalanda (n = 5,728, 70.1%) than in Saran (n = 2,446, 29.9%).

**Fig 2:**
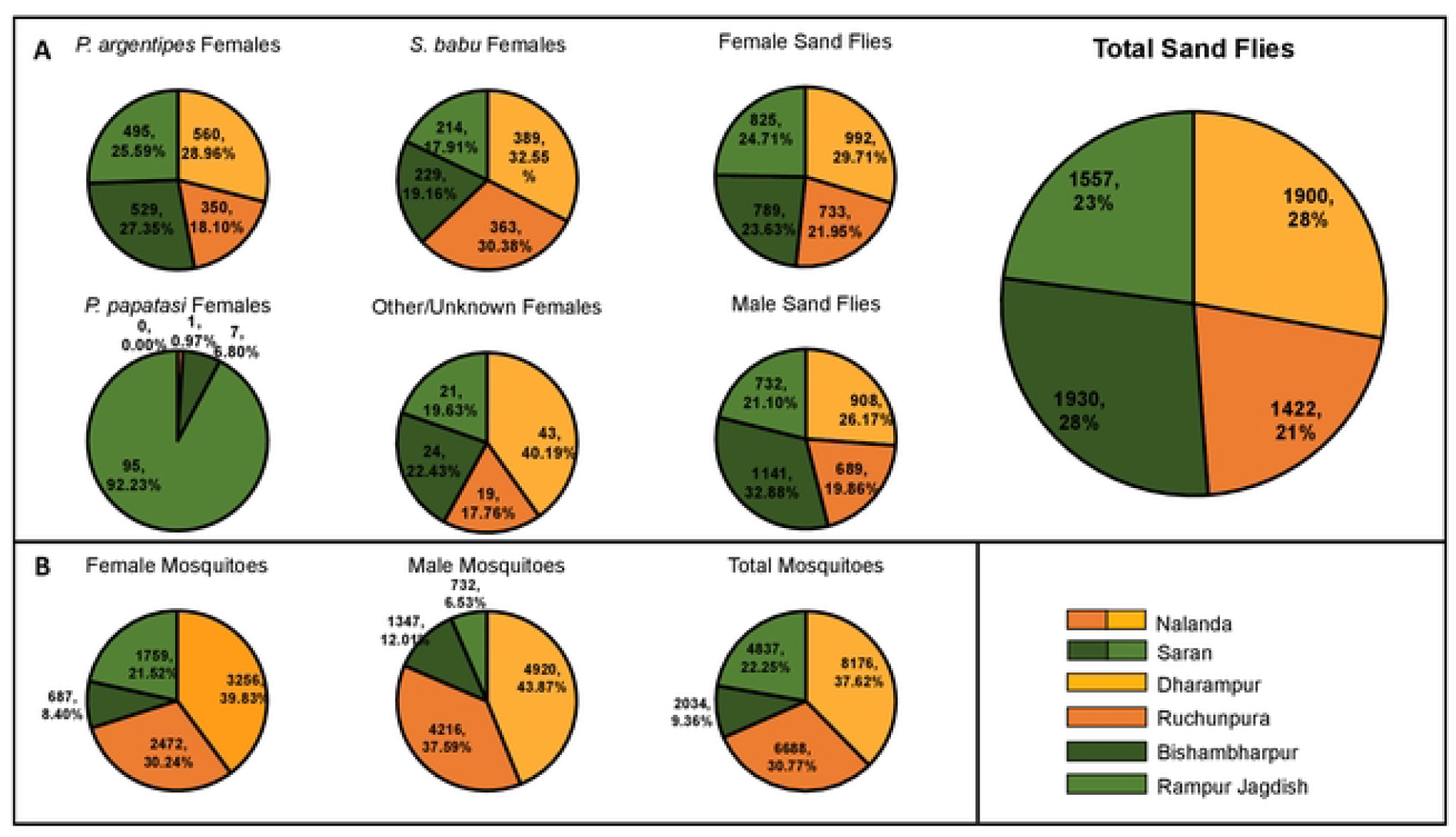
Specimens collected by sex and collection location over the study period. (A) displays sand fly data, (B) displays mosquito data.

#### Collection method

The total numbers of specimens by collection method for all four rounds is given in supplementary table S1. Fig 3 shows the mean number of sandflies collected by district, village and by collection method. CDC-LTs collected the highest mean number of sandflies compared to MVA and PKP in both districts. CDC-LTs accounted for 57.5% of total sand flies collected, followed by PKP and MVA with 22.4% and 20.1%, respectively.

**Fig 3:**
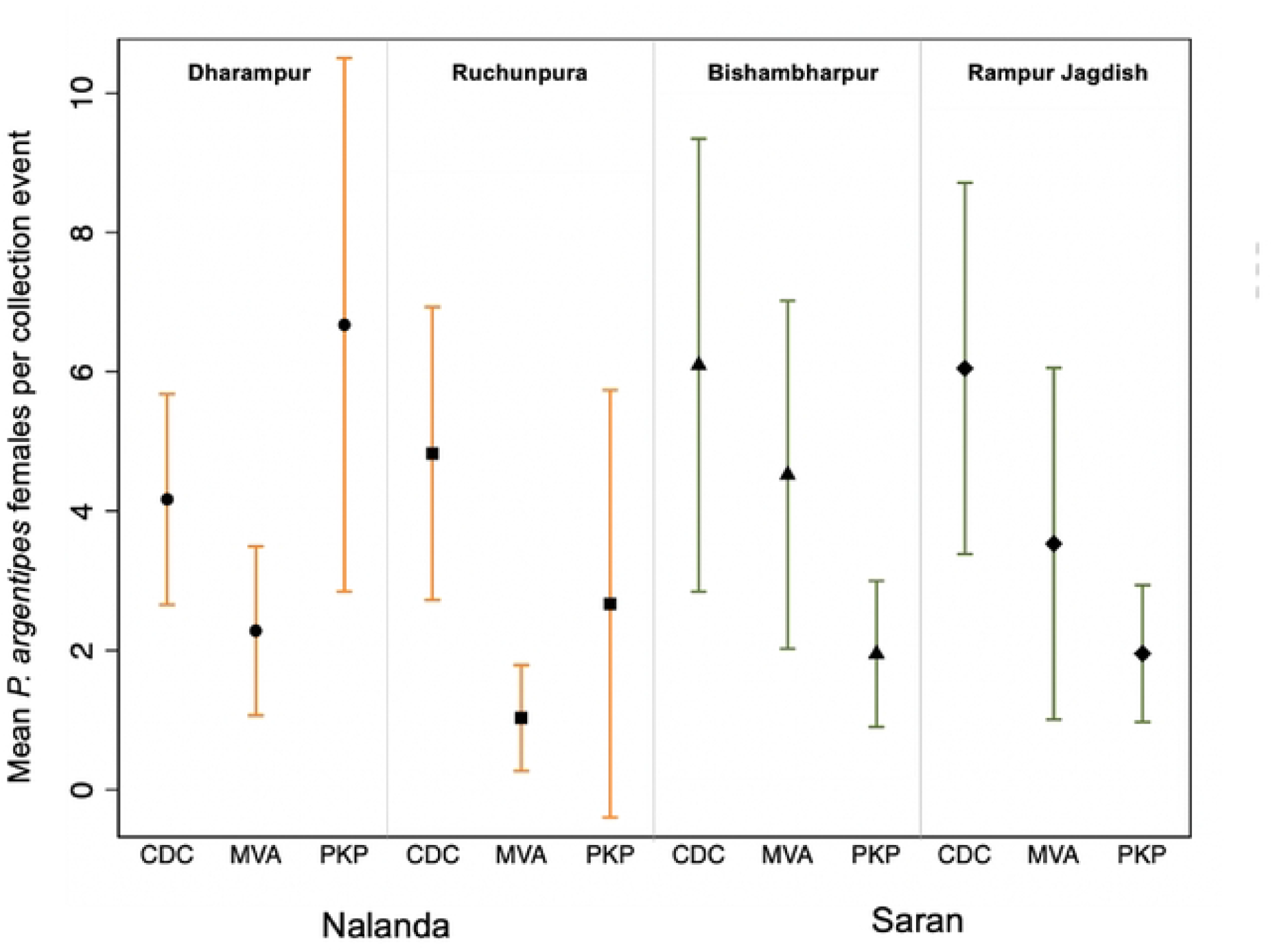
Mean female sand flies collected by method and village. Error bars indicate the 95% confidence interval of the mean.

Fig 4 shows the mean number of female mosquitoes collected by district, village and by collection method. Bishambharpur had the lowest mean number of mosquitoes for all three collection methods. Collections in Dharampur using CDC-LTs had the highest overall mean female mosquitoes by village and method (32.5). The overall mean number of female mosquitoes per collection event was 14.5; CDC-LT yielded the greatest mean per collection event (16.96) while PKP yielded the lowest (10.0).

**Fig 4:**
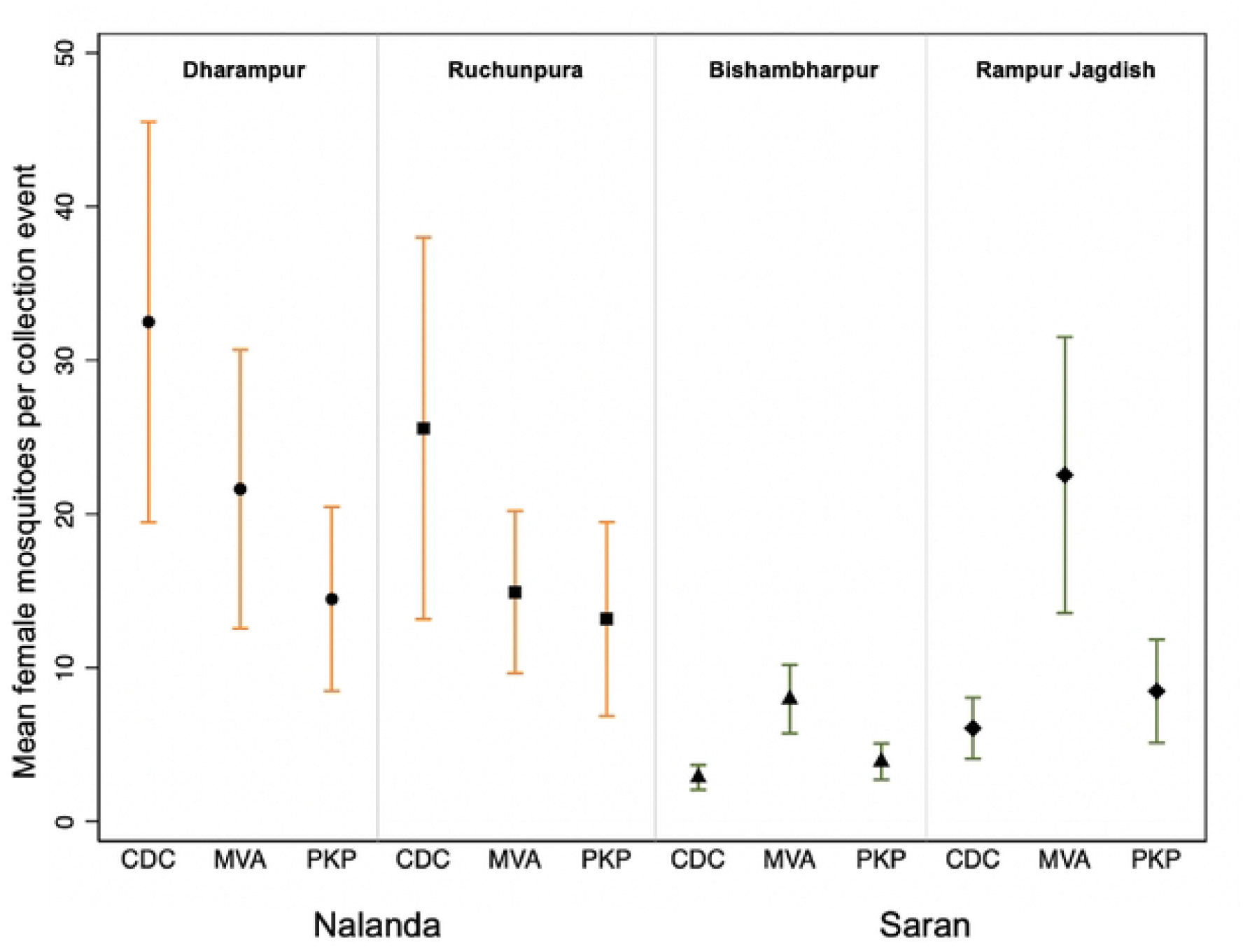
Mean female mosquitoes collected by method and village. Error bars indicate the 95% confidence interval of the mean.

Further descriptive and regression analyses focused on *P. argentipes* females in the sand fly collections and were limited to the first three collection rounds due to post-collection loss of sand fly specimens from round four.

### Statistical analyses for rounds 1-3 of CMC

The total numbers of specimens by round and village, and by collection method, for the first three rounds of collections are given in supplementary tables S2 and S3, respectively. Sampling round 2 yielded the highest mean for total sand flies, male sand flies, and female sand flies, with mean collections double those of round 1. Round 3 yielded the highest mean *P. argentipes* (Fig 5), with almost 2.5 times the collection of round 1, but the total mean sand flies collected was lower in round 3 than for round 2 for both males and females.

**Fig 5:**
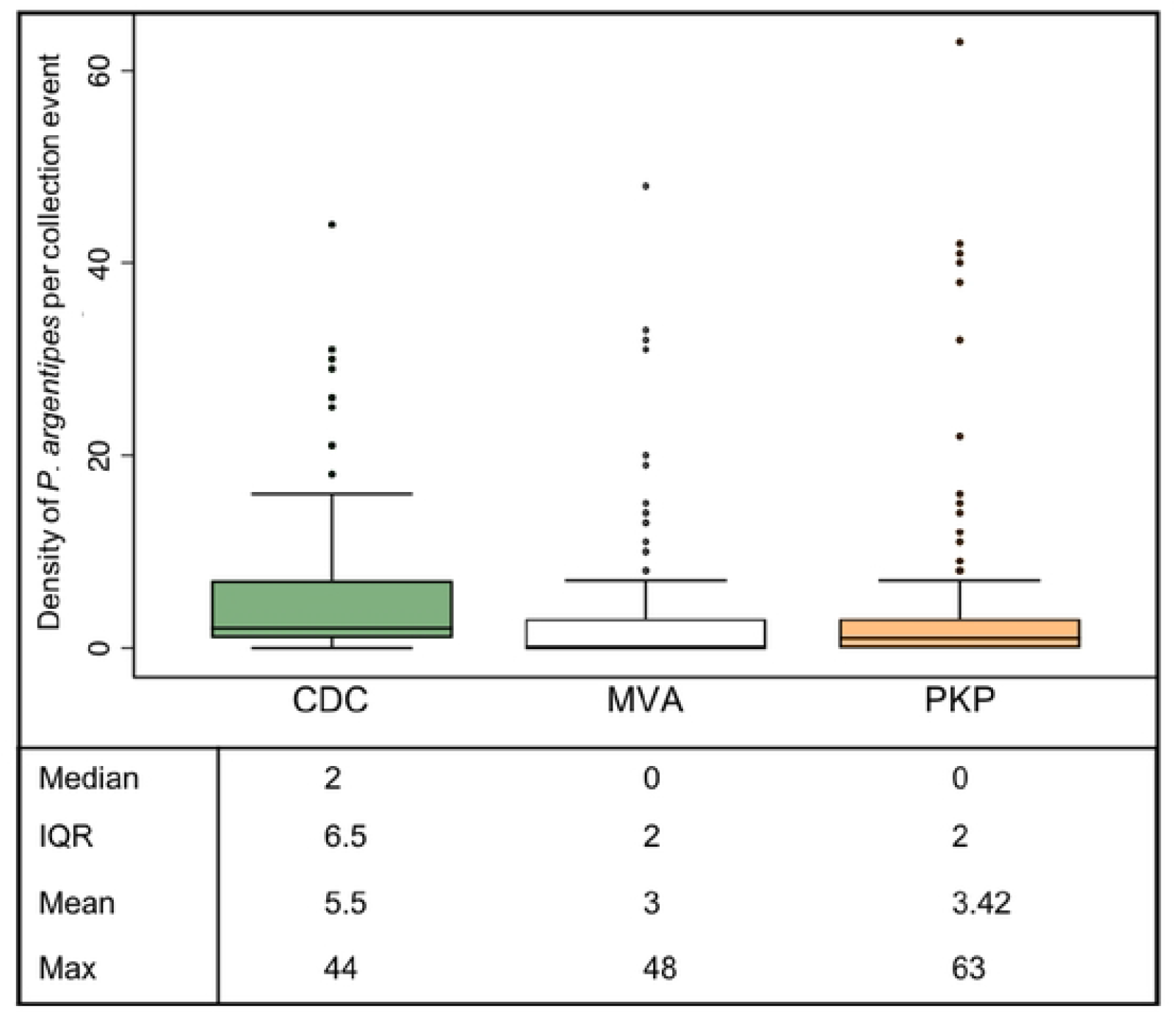
Box plots showing the number of *P. argentipes* females/collection event by method of collection. Medians indicated by horizontal line (at 0 for MVA and PKP) and outliers by black dots.

Logistic regression analysis comparing detection (presence) of sand flies, female sand flies, and female *P. argentipes* showed CDC-LTs were significantly different from both MVA and PKP with increased odds of detection from CDC-LT collection events (p<0.0001). Of 511 collection events (excluding missing data), 75.6% (124/164) CDC-LTs collections resulted in the capture of at least one female *P. argentipes* compared to 43.2% (76/176) for MVA and 56.2% for PKP (87/171). Fig 6 shows the density of female *P. argentipes* by collection method. CDC-LTs captured the highest density of female *P. argentipes* per trap-night compared with MVA or PKP. However, the greatest number of female *P. argentipes* from a single collection event (n=63) was collected by PKP. After adjusting for household clustering, CDC-LTs collected 3.41 times more female sand flies, 4.04 times more *P. argentipes*, and 8.23 more male sand flies than MVA (p<0.0001 for each). CDC-LTs also collected 3.08 times more female sand flies, 3.62 times more *P. argentipes*, and 7.21 more male sand flies than PKP (p<0.0001 for each) (Table 1). PKP collected significantly more female mosquitoes than MVA (p = 0.0002), but there were no significant differences between CDC-LTs and PKP or MVA.

**Table 1:** Number of female sand flies, male sand flies, *P. argentipes* females, and female mosquitoes by collection method including IRR and negative binomial regression analyses.

|  |  | N | IRR (95% CI) | Z | P-value |
| --- | --- | --- | --- | --- | --- |
| Total Female Sand Flies |  |  |  |  |  |
|  | MVA | 685 | 0.293 (0.215 – 0.401) | -7.70 | <0.0001 |
|  | PKP | 745 | 0.325 (0.238 – 0.442) | -7.13 | <0.0001 |
|  | CDC | 1441 | 1 | - | - |
| *P. argentipes* Females |  |  |  |  |  |
|  | MVA | 426 | 0.247 (0.161 – 0.380) | -6.39 | <0.0001 |
|  | PKP | 468 | 0.276 (0.181 – 0.421) | 0.51 | <0.0001 |
|  | CDC | 726 | 1 | - | - |
| Male Sand Flies |  |  |  |  |  |
|  | MVA | 371 | 0.121 (0.086 – 0.171) | -12.13 | <0.0001 |
|  | PKP | 435 | 0.138 (0.099 – 0.194) | -11.46 | <0.0001 |
|  | CDC | 2019 | 1 | - | - |
| Female Mosquitoes |  |  |  |  |  |
|  | MVA | 2097 | 1.22 (0.915 – 1.648) | 1.37 | 0.172 |
|  | PKP | 1440 | 0.791 (0.595 – 1.053) | -1.60 | 0.109 |
|  | CDC | 1843 | 1 | - | - |

**Fig 6:**
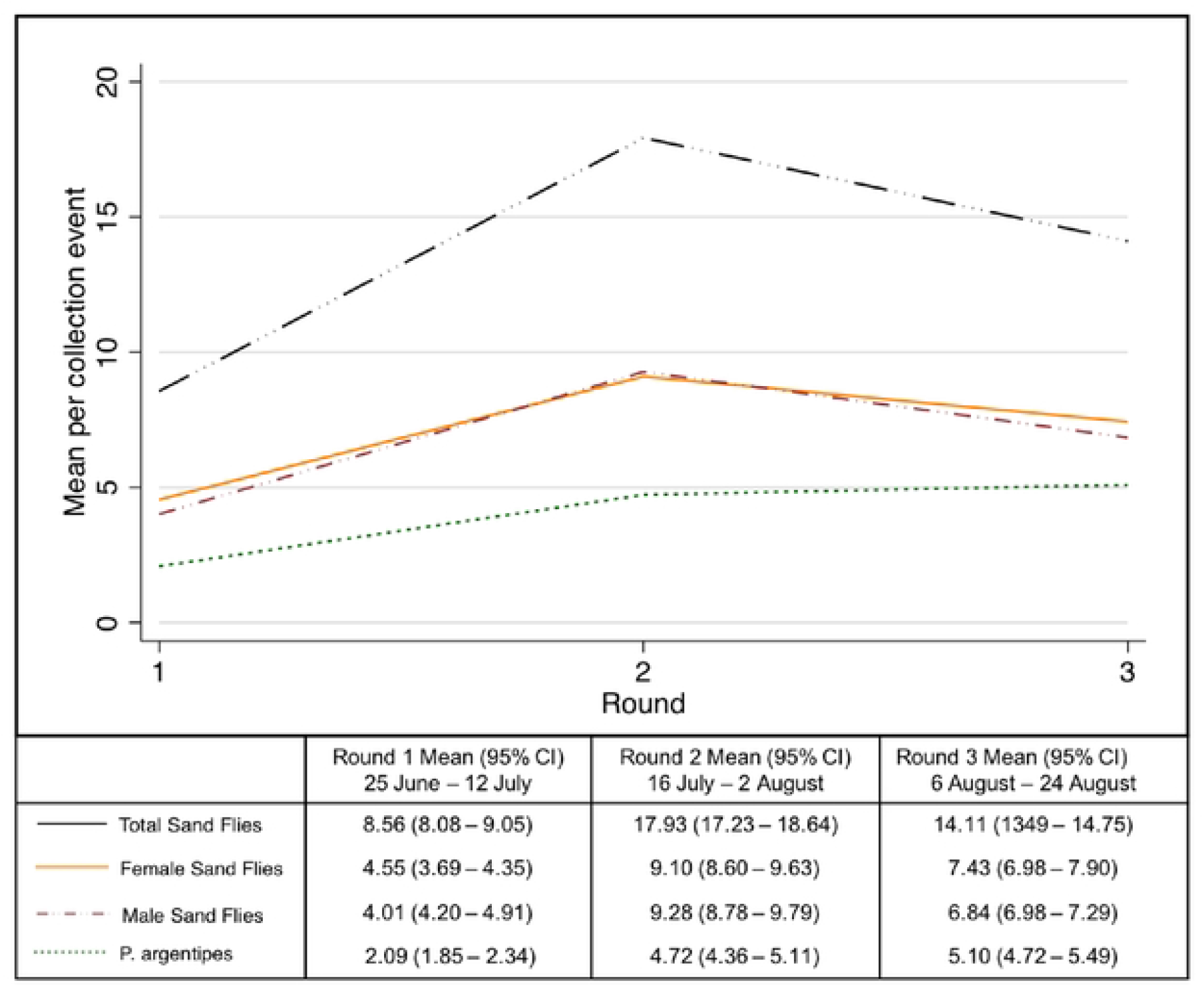
Mean sand flies collected per trap-night by collection round for the first three collection rounds (with 95% CI).

### Molecular Analyses

#### Detection of human and Leishmania donovani DNA

Of 1,934 female *P. argentipes*, 1,932 were analysed for the presence of a human DNA derived from a bloodmeal, and 156 (8.1%) were positive. Almost 75% of *P. argentipes* with human DNA were collected from the two Saran villages. As a proportion of total female *P. argentipes* caught, MVA collected significantly more female sand flies with human DNA compared with CDC-LTs and PKP (12.97% versus 5.65% and 7.40%, respectively, p-value<0.003).

Of 1,934 *P. argentipes* analysed for the presence of *L. donovani* DNA, none were positive.

#### Simulation of Leishmania donovani RNA degradation

When the infection rate was low there was an increase in the number of cycles for amplification (Fig 7). However, even with low infection rates, most samples were amplified before 30 cycles indicating good sensitivity. For the ‘worst case scenario’, simulating CDC-LT samples where sand flies were collected overnight for 16h at 40°C and left in a cool bag for 7 hours, *L. donovani* RNA was detected in sand flies with results comparable with conditions simulating aspirator collections.

**Fig 7:**
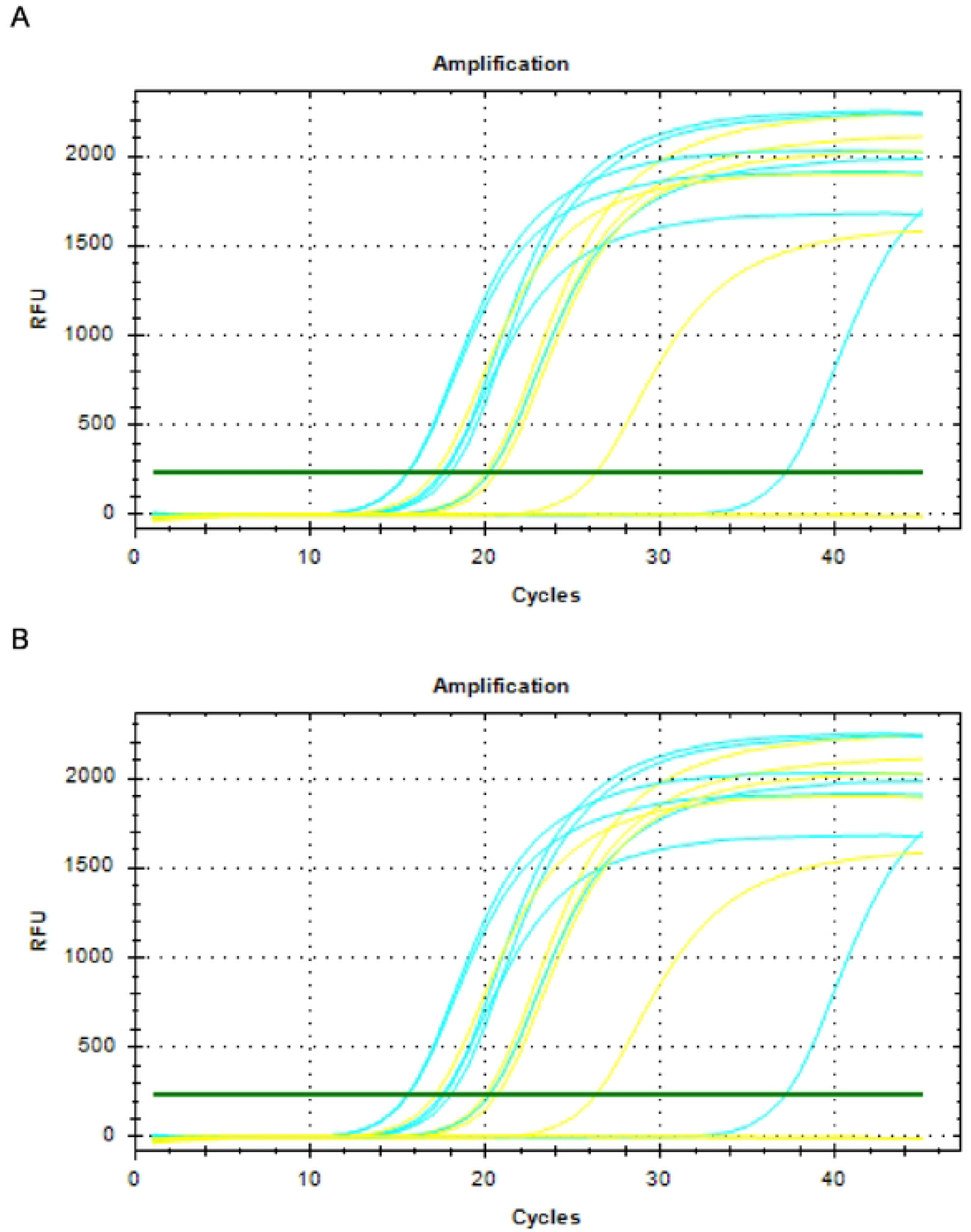
Amplification curves for *Leishmania donovani* in sand flies at (A) high (3000 amastigotes/sandfly) or (B) low (<100 amastigotes/sandfly) infection rates maintained under simulated conditions for aspirator (blue) or CDC collections (yellow) and kept in a cool bag for 7 hours.

### Operational considerations

#### Time burden of sampling

On average, CDC-LTs took less time to install and to collect (6.29 mins) than the time taken to perform aspirations when using either MVA or PKP by 10.14 and 12.85 mins, respectively. The time taken to sort samples was influenced by the amount of material and number of target and non-target specimens within the collections. Although the mean time taken to sort CDC-LT collections (12.50 mins) was longer than for collections obtained using MVA or PKP by 1.29 and 7.19 mins, respectively, they also contained the highest number of female sand flies (n=3), male sand flies (n=6) and female mosquitoes (n=157) than MVA (n=0,1,92) or PKP (n=1,0,3), respectively. Therefore, proportionally, the time invested in sorting compensated for the yield obtained.

#### Householder acceptance

Responses were received from all 48 householders who took part in the study. Amongst the respondents, 36 (75%) indicated they had a preference for CDC-LTs, 6 (13%) for MVA, 2 (4%) for PKP/MVA, and 4 (8%) had no preference. No householder indicated they had a preference solely for PKP collections. The most common reason given for a preference for CDC-LTs was the perception that fewer mosquitoes were present in their household following sampling. However, it is interesting to note that amongst householders who indicated a preference for either MVA or MVA/PKP, the most common explanation was also a perception that there were fewer mosquitoes present in the household after sampling. Three householders indicated that they felt CDC-LTs were the easiest collection technique, one considered it to be a less disruptive method, and two were unable to justify their choice. One respondent preferred MVA collections because they felt they had insufficient space in their home for a CDC-LT to be installed. When prompted for additional comments or complaints about the collection techniques, five householders responded that they found CDC-LTs to be too noisy at night and this caused disruption to their cattle. No further comments or complaints were received for either the PKP or MVA.

## Discussion

This is the first comprehensive field study that has made a direct comparison between CDC-LTs and two types of large battery-powered aspirators used in mosquito surveillance for the capture of *P. argentipes* females. Traditionally, CDC-LTs and mouth aspirators, rather than large battery-powered aspirators, are used in field studies performed in India to capture *P. argentipes* females (11, 17, 21, 26). However, a decision was made to avoid using mouth aspirators in the present study due to health and scientific concerns. Firstly, even if mouth aspirators are equipped with filters, the potential risk to the field team when making repeated aspirations over a long period of time, inevitably involving ingestion of small particles of organic debris while gently sucking sand flies from walls in a tube, was considered too high. Secondly, the degree of potential bias in efficiency between and amongst collectors, as well as problems relating to standardisation of the technique, was inappropriate for comparing collection methods. Instead, two different battery-powered aspirators were compared using a standard operating procedure that was developed for the study.

### Collection Method Comparison Study

As expected, capture rates were highly variable given they are affected by several factors, including site, seasonality and climatic variables, which highlights the importance for testing the different collection methods at the same sites and within the same timeframe, and reducing sampling bias by using a Latin Square design, as performed in the present study. It is possible that capture rates using either type of aspirator may have been higher if collections were performed at an earlier or later time of the day than the time window of 07.00 to 14.00 selected in the present study which was consistent with working hours of operational field teams rather than those that may be ideal for research purposes.

Nevertheless, it is worth noting that the mean capture rates of *P. argentipes* females/trap night using CDC-LTs placed in human dwellings, of 4.2-4.8 or 6.0-6.1 *P. argentipes* females/trap night in Nalanda and Saran villages, respectively, were consistent with a mean capture rate of 5.5 *P. argentipes* females/trap night found in a previous study performed in Saran, but higher than the mean capture rate of 2.0 *P. argentipes* females/trap night reported in Muzaffarpur, Bihar, another VL endemic district bordering Saran (11, 21) and in other sentinel sites in India where, even during the June-September peak, densities of *P. argentipes* females/trap night declined from 2014-2019 (27).

A complete data set, comprising speciation of *P. argentipes*, human DNA detection and *L. donovani* DNA detection, was available for three rounds (three 12×12 Latin squares) of collections (n=144 collection events/method), rather than for all four rounds performed, and these data were used in statistical analyses. Significant differences between collection methods were detected for the primary outcome, number of *P. argentipes* females, since the study was still sufficiently powered to detect a mean difference of between 1.00-1.25 *P. argentipes* females using the data used to perform the sample size calculation (18, 21). The main finding that CDC-LTs collected an average of either 4.04 times or 3.62 times more *P. argentipes* females than either of the battery-powered aspirators, MVA or PKP, respectively, supports their continued use in entomological surveillance but other factors should be considered as described further below. It is interesting to note that, for the purposes of integrated vector-borne disease surveillance and multiplex pathogen screening, there were no significant differences between incidental captures of female mosquitoes using CDC-LTs compared with PKP or MVA.

To determine which human pathogens are circulating within an area, some MX studies use collection methods that specifically target blood-fed females to screen for infection. For studies measuring transmission, when it is important to maximise the likelihood of obtaining vectors infected with the human infectious stage of the parasite, then collection methods that target gravid vectors are used. Methods that target host-seeking females, such as CDC-LTs, capture unfed females which may be nulliparous (never blood-fed so will not be infected unless the pathogen is transovarially/transstadially transmitted) or parous (previously blood-fed, has laid eggs and seeking next bloodmeal so may also be infected/infectious). Some research groups include parity as an entomological outcome in their studies, but parity was not examined in the present study because it is time consuming, can be misclassified according to expertise, and is unlikely to be incorporated routinely in programmatic use.

### Molecular analyses

Even though CDC-LTs collected proportionally less *P. argentipes* females with human DNA than MVA, there was no significant difference in the absolute number of *P. argentipes* females with human DNA captured. In the present study, the mean rate of human DNA detected in *P. argentipes* females for all three collection methods was 8.1% with a mean rate of 5.4% for CDC traps placed inside houses. This rate is slightly higher than previously reported in Saran, where bloodmeals were detected in 3.2% and 2.6% of *P. argentipes* collected in CDC-LTs placed inside combined dwellings (where humans and cattle are in close proximity) and houses (with no cattle), respectively (11). Previously, in Muzaffarpur, the mean percentage of blood-fed *P. argentipes* captured in CDC-LTs was 30.2% pre-intervention and 2.4% post-intervention with untreated bed nets (28). Although it is possible that low feeding rates in Saran may be due to IRS intervention, there was no significant difference between the mean numbers of *P. argentipes* collected in households between Saran (VL endemic) and Nalanda (non-programmatic) villages, and the mean rate of human DNA detected in *P. argentipes* collected in Saran villages (11.4%) was significantly higher than for the non-endemic Nalanda villages (4.4%). There are several factors that may account for the differences, including: house construction (influencing sand fly entry and/or providing optimal conditions for resting), number of human inhabitants/house, type/number of livestock (either providing protection or increasing the risk of biting), and implementation of IRS (excito-repellency effect of synthetic pyrethroids may increase the risk of outdoor biting).

Human DNA detected in the sand flies could derive from a bloodmeal taken solely from a human host, or from a sand fly that had fed on multiple hosts since mixed bloodmeals have been reported for *P. argentipes* captured previously in Saran where human DNA was detected using cyt *b* PCR and reverse line blot in 164 of 288 (57%) blood-fed females (13). The same study showed that the peak time to detect human DNA in *P. argentipes* in Saran villages was June-September which coincided with the sampling period performed in the present study (13). In Muzaffarpur, the Human Bloodmeal Index was 81% pre-intervention and 19% post-intervention with untreated bed nets (28).

There are few studies measuring infection rates of *P. argentipes* with *L. donovani* DNA in India. Previously, CDC-LT collections performed in 2010 in Muzaffarpur showed that infection rates vary from 0.9% to 2.8% (12/422) according to the season (16). In the present study, no *L. donovani* DNA was detected in each of the 1,934 sand flies individually tested from collections performed in 2018 (0/1024 from Saran and 0/910 from Nalanda). The inability to detect *L. donovani* DNA in *P. argentipes* in the Saran villages is surprising considering that VL and PKDL cases were reported on the KAMIS system in months leading up to and after when the work was performed. However, similar low infections rates in *P. argentipes* were also reported in eight VL sentinel sites undergoing routine entomological and IRS surveillance in three VL endemic States in India (27). Of 14,775 *P. argentipes* females tested, *L. donovani* DNA was detected in only four females collected in the district of East Champaran, Bihar (3/991 and 1/704 females collected 2017 and 2018, respectively). No females were positive at any other of the sentinel sites suggesting that active transmission is low (27). These data provide further evidence to support that efforts implemented during the accelerated kala-azar control programme may have successfully reduced the VL burden in endemic blocks.

It is important to note that differences in the sensitivity and specificity of the molecular assays used may affect differences in results. Many assays have been developed with different molecular targets with a specificity variation between 29.6–100% and sensitivity between 91.3–100% has been reported (25). In addition, most assays used were developed to detect infections in human patients or dogs, rather than sand flies, and may cross-react with common insect protozoan parasites of the family Trypanosomatidae (e.g. *Leptomonas*). To avoid false positive or false negative results, it is important to use assays with targets specific for *L. donovani* and also use more than one target to confirm results, as performed in this study.

Also crucial for the purposes of MX, was the finding from the simulation experiment that the higher yield of *P. argentipes* females in CDC-LTs was unlikely to be compromised by an inability to detect *Leishmania* RNA in samples since RNA analysis may be required in future studies aiming to detect the metacyclic infectious stage in sand flies.

### Operational considerations

Firstly, it should be noted that battery procurement and availability can be a limiting factor for all three methods used. It took a substantial effort to obtain the three different types of recommended batteries for the collection methods in India, but a supplier for large numbers of 6V 12amp batteries, as used for CDC-LTs, was later identified. Further evidence in favour for using CDC-LTs was the lower time burden for collecting specimens than either method of aspiration. This finding has some major implications for operational teams since it influences how many households can be sampled in one day: roughly 20 households may be sampled/day using CDC-LTs (including setting up and collecting traps) for every 9-10 or 10-11 aspirations using PKP or MVA, respectively. Sorting samples, and accurate recording of data, in the laboratory needs to be performed with precision and, although it took less time to sort samples collected using PKP than for the other methods of collection in our supplementary study, they contained less target material. It should be noted that the supplementary study performed to obtain time estimates took place in February, when sand fly numbers tend to be lower than June to September when the CMC study took place (11). However, results of the CMC indicate that although collections using aspirators will contain less sand flies to process than collections using CDC-LTs, they will have similar numbers of female mosquitoes during the June to September peak and mosquitoes outnumber sand flies in the collections.

Importantly, more householders in the study expressed a preference for CDC-LTs than for the battery-powered aspirators in contrast to residents in Brazil who preferred MVA over CDC-LTs due to the nuisance of light and noise in the bedroom at night (8). Continued consent of householders in India to allow indoor sampling using CDC-LTs in their homes should be monitored especially in light of some complaints relating to disturbance to animals expressed in the current study.

### Recommendations

Based on the evidence obtained, CDC-LTs are the best method for collecting *P. argentipes* females in houses for transmission studies. However, capture rates are still low, and further research is required to increase collections of target species and to improve molecular methods. Specifically, in order to calculate the EIR of *P. argentipes*, a diagnostic test must be developed to identify the metacyclic stage of *L. donovani*. Although a laboratory protocol for detecting *L. mexicana* metacyclics in infected *Lutzomyia longipalpis* sand flies has been published (29), it has not yet been validated in the field nor modified for other *Leishmania*/sand fly models.

Presently, molecular methods used for MX require skilled technical staff and a well-equipped laboratory and the cost of screening large numbers of vectors may be prohibitive for programmatic use thus reducing the role of MX for research purposes only (6). To expand the flexibility of a MX system for programmatic use, and make it more cost-effective, integrating vector-borne disease surveillance should be considered. New multiplex methods for point-of-need use are under development which make the possibility of integrating vector-borne disease surveillance a realistic and cost-effective proposition.

## Data Availability

All data will be made available.

## Acknowledgements

The research team would like to thank the following individuals and organizations: All householders in Nalanda and Saran who permitted access to their homes.

HMSC for providing permission for the study. The National Centre for Vector Borne Disease Control (formerly the Vector Borne Disease Control Programme) for their support and advice in placement of the research.

Field workers and project administration staff who participated in this research. Dr Tom Walker for advice on molecular tests.

## CRediT authorship contribution statement

**Shannon McIntyre-Nolan:** Conceptualization, Investigation, Methodology, Writing - Original Draft Preparation, Writing - Review & Editing. **Vijay Kumar:** Project Administration, Resources, Writing - Review & Editing. **Miguella Mark Carew:** Data Curation, Formal Analysis, Writing - Original Draft Preparation, Writing - Review & Editing. **Kundan Kumar:** Investigation, Writing - Review & Editing. **Emily Nightingale:** Formal Analysis, Methodology, Writing - Original Draft Preparation, Writing - Review & Editing. **Giorgia Dalla Libera Marchiori:** Investigation, Methodology, Writing - Review & Editing. **Matthew Rogers:** Investigation, Supervision, Writing - Review & Editing. **Mojca Kristan:** Investigation, Writing - Review & Editing. **Susana Campino:** Investigation, Supervision, Methodology, Validation, Writing - Original Draft Preparation Writing - Review & Editing. **Graham F. Medley:** Formal Analysis, Funding Acquisition, Supervision, Writing - Review & Editing. **Pradeep Das:** Conceptualization, Resources, Supervision, Writing - Review & Editing. **Mary Cameron:** Conceptualization, Funding Acquisition, Methodology, Project Administration, Supervision, Writing - Original Draft Preparation, Writing - Review & Editing.

## Informed Consent, Confidentiality, and Ethical Approval

The study was approved by the Ethics committee of the GOI Health Ministry’s Screening Committee (Ref: 2017-4126), RMRIMS (Ref: 39/RMRI/EC/2017) and the London School of Hygiene & Tropical Medicine (Ref: 1463). Members of each household were provided with an information sheet in Hindi explaining the purpose of the study, their involvement, and any potential adverse outcomes prior to commencement of sampling at each site. Information sheets were read to participants if they identified themselves as illiterate. Participants met with the investigators to answer any questions about the study. The household head or a nominated proxy provided written or oral informed consent for recruitment to the study. Signed consent forms and audio consent recordings were stored securely to protect participant confidentiality.

## Supplemental Tables

**S1.** Specimens collected by collection method for all four rounds of the study.

| Rounds1-4 | Collection Method |  |  | Total |
| --- | --- | --- | --- | --- |
|  | CDC | MVA | PKP |  |
| **Total sand fly females** | 1621 (48.5) | 810 (24.3) | 908 (27.0) | **3,339** |
| *P. argentipes* females | 866 (44.8) | 501 (25.9) | 567 (29.3) | **1,934** |
| *P. papatasi* females | 24 (23.3) | 47 (45.6) | 32 (31.1) | **103** |
| *Sergentomyia babu* females | 704 (58.9) | 224 (18.7) | 267 (22.3) | **1195** |
| Other/Unknown females | 27 (25.2) | 38 (35.5) | 42 (39.3) | **107** |
| **Total sand fly males** | 2293 (66.1) | 558 (16.1) | 619 (17.8) | **3,470** |
| **Total sand flies** | 3914 (57.5) | 1368 (20.1) | 1527 (22.4) | **6,809** |
| **Total female mosquitoes** | 3085 (37.7) | 3193 (39.1) | 1896 (23.2) | **8174** |

**S2.**
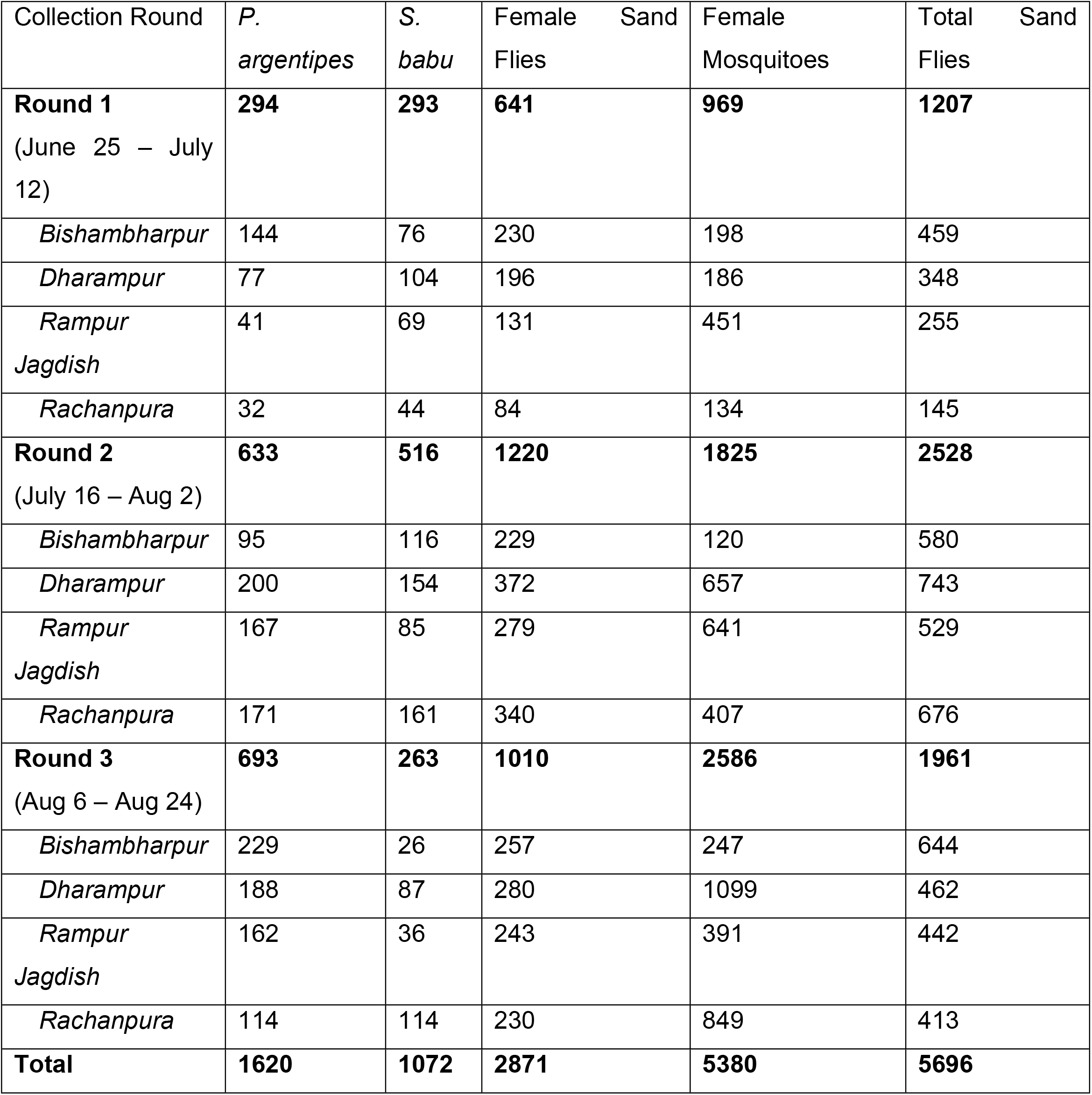
Specimens collected by round and village.

**S3.** Numbers and percentages of sandflies and mosquitoes collected by species, sex, and collection method during the first three collection rounds used in logistic regression analysis.

| Rounds 1-3 | Collection Method |  |  | Total |
| --- | --- | --- | --- | --- |
|  | CDC-LT | MVA | PKP |  |
| **Total sand fly females** | 1441 (50.2) | 685 (23.9) | 745 (25.9) | **2871** |
| *P. argentipes* females | 726 (44.8) | 426 (26.3) | 468 (28.9) | **1620** |
| *P. papatasi* females | 24 (28.2) | 34 (40.0) | 85 (31.8) | **85** |
| *Sergentomyia babu* females | 664 (61.9) | 187 (17.4) | 221 (20.6) | **1072** |
| Other/Unknown females | 27 (28.7) | 38 (40.4) | 29 (30.9) | **94** |
| **Total sand fly males** | 2019 (71.5) | 371 (13.1) | 435 (15.4) | **2825** |
| **Total sand flies** | 3460 (60.7) | 1056 (18.5) | 1180 (20.7) | **5696** |
| **Total female mosquitoes** | 1843 (34.3) | 2097 (39.0) | 1140 (26.8) | **5380** |

